# Projected impact of a national Tobacco 21 policy in the Kingdom of Saudi Arabia

**DOI:** 10.64898/2026.02.25.26347109

**Authors:** Jamie Tam, Rafael Meza, Mohammed Abdullah Aljabri, Abdulmohsen H. Al-Zalabani, Sarah S. Monshi, Ahmed Ali Yakoub, Fatmah Mohammed Aldhaher, Mariam M. Hamza, Wafi Albalawi, Reem Alsukait, Mohammed Adeeb Shahin, Volkan Cetinkaya, Taghred Alghaith

## Abstract

**Introduction:** Tobacco smoking is especially high among males in the Kingdom of Saudi Arabia (KSA). In 2019, 27.5% of males ages 15+ reported smoking. Despite a minimum age requirement of 18 years, data indicate that 6.8% of youth ages 13-15 currently smoke tobacco products. To reduce youth smoking, countries have raised the minimum purchase age to 21, also referred to as Tobacco 21. Except for Kuwait, no other Middle Eastern country has done so. We develop a tobacco smoking simulation model to project the potential impact of a national Tobacco 21 policy in Saudi Arabia.

**Methods:** We used data from three nationally representative health surveys in Saudi Arabia to develop the KSA Tobacco Control Policy (TCP) model, estimating smoking initiation and cessation rates for males, as smoking rates are low among females. A national Tobacco 21 policy was operationalized as a 34% (15%-53%) reduction to smoking initiation for ages 18-20. Economic impact was evaluated using the 2024 KSA value of a statistical life which ranges from $1.65 million to $5.15 million USD.

**Results:** Under a status quo scenario, tobacco smoking prevalence in males would decrease to 10.2% by 2100. Implementation of Tobacco 21 in 2026 would decrease smoking prevalence to 9.4% (8.9%, 9.8%) by 2100. While modest, these reductions would eventually translate into nearly 5000 (2200, 7800) premature deaths averted with up to 155000 (69000, 241000) life years gained from 2026-2100, respectively. The total expected economic benefit ranges from $1.67 to $5.19 billion USD, equivalent to 6.25 to 19.45 billion SAR.

**Discussion:** Timely implementation would support the KSA in its goals to reduce non-communicable disease and death; however, even under best-case conditions, a Tobacco 21 alone would not achieve the Vision 2030 smoking prevalence target of 9%. Additional policies that substantially increase smoking cessation are needed.

**What is already known on this topic:** The leading causes of death in Saudi Arabia are all linked to tobacco smoking. Tobacco 21 policies have been pursued by numerous governments to reduce youth smoking, but such policies are lacking in Middle Eastern nations.

**What this study adds:** A nationwide Tobacco 21 policy in Saudi Arabia would reduce smoking initiation, smoking prevalence, and smoking-related mortality. Overall smoking prevalence among males ages 15+ would decline, and nearly 5000 premature deaths would be averted with up to 155,000 life years gained from 2026-2100, valued at 6.25 to 19.45 billion SAR.

**How this study might affect research, practice or policy:** This study quantified for the first time the potential long-term benefits of a Tobacco 21 policy in Saudi Arabia for the male population. A Tobacco 21 policy would benefit future generations of young people by reducing their risk for heart disease, stroke, and cancer, currently the leading causes of death in the nation. However, additional efforts are needed in addition to Tobacco 21 policies to achieve tobacco smoking reduction goals.

## INTRODUCTION

In the Kingdom of Saudi Arabia (KSA), the top three leading causes of death are ischemic heart disease, stroke, and cancer—all of which are caused by smoking.^1^ The most commonly used tobacco products are cigarettes and waterpipe (shisha), though other products such as e-cigarettes and nicotine pouches have also entered the market. Data from the 2019 Global Adult Tobacco Survey (GATS) shows that nearly 1 in 5 adults ages 15 or older uses tobacco, with much higher prevalence among men (30%) compared to women (4%).^2^ Among youth, 1 in 10 boys and 1 in 12 girls reports tobacco use.^3^ Alarmingly, the proportion of youth who initiated cigarette smoking at age 7 or younger nearly doubled from 2007 (9.1%) to 2022 (17.1%).^4^ These data highlight the need for progressive tobacco control measures that have the greatest potential to reduce the non-communicable disease burden, improve overall health, and extend life expectancy in the KSA.

The Kingdom of Saudi Arabia has shown its commitment to addressing tobacco use through policy action.^5^ Saudi Arabia ratified the WHO Framework Convention on Tobacco Control in 2005, and has since banned most forms of advertising and promotion of tobacco products, required plain packaging and graphic health warnings on cigarette packs, and prohibited indoor smoking in most workplaces and public facilities.^6^ These progressive public health actions made the country a global leader in tobacco control, as evidenced by the awarding of the 2019 WHO Tobacco Control Medal to Dr. Tawfig bin Fawzan Al-Rabiah, the then Saudi Minister of Health, for the Ministry’s leadership in this arena.^7^ Under Saudi Arabia’s strategic plan for tobacco control, leaders aim to reduce smoking to 9% by 2030.^8^ Currently, the sale of tobacco products is prohibited to anyone under age 18 in the country, yet data from the 2022 Global Youth Tobacco Survey indicate that 6.8% of youth ages 13-15 currently smoke tobacco products, including shisha.^3^

To address smoking among young people and protect future generations from the harms of tobacco, countries around the world have raised the minimum purchase age to 21, also referred to as Tobacco 21. Currently, Kuwait is the only country in the Middle East with a Tobacco 21 policy.^9^ However, some countries in other regions have implemented national Tobacco 21 policies, including Ethiopia, Honduras, Kazakhstan, Singapore, Sri Lanka, Uganda, Uzbekistan, and the United States (US).^10,11^ To our knowledge, specific evidence on the effects of Tobacco 21 for most countries is not available, except from the US. Research from the US shows that Tobacco 21 policies reduce smoking among young people.^12^ Simulation modeling analyses estimate that the expected reductions in smoking due to Tobacco 21 policies in the US will result in substantial mortality reductions and life-years gained.^13^ As Saudi Arabia works toward its goal of reducing smoking and smoking-related disease and death, information about the likely public health benefits of a national Tobacco 21 policy is needed.

The primary objective of this study is to develop a simulation model of smoking that can evaluate the potential long-term impacts of a national Tobacco 21 policy in the Kingdom of Saudi Arabia. Findings can inform decision-making regarding tobacco control policies for the population and quantify the expected progress if a Tobacco 21 policy is implemented.

## METHODS

### Data sources

To develop a model of smoking for Saudi Arabia, we used data from the 2013 Saudi Health Interview Survey (SHIS, n=10,821), the 2019 Global Adult Tobacco Survey (GATS, n=11,381), and the 2019 World Health Survey (WHS, n=8912). Data on cigarette smoking initiation and cessation are not disaggregated from tobacco smoking, so our definition of smoking includes other non-cigarette forms of smoked tobacco, such as waterpipe or shisha. SHIS is a nationally representative household survey that collected health behavior and disease information among adults aged 15 or older, which was conducted by the Saudi Arabia Ministry of Health and the Institute for Health Metrics and Evaluation of the University of Washington.^14^ We used SHIS individual data on self-reported current, former, and never tobacco smoking prevalence as well as data on the age at daily tobacco smoking initiation. GATS is a nationally representative household survey of adults aged 15 or older, part of the Global Tobacco Surveillance System led by the US Centers for Disease Control and Prevention. We used GATS individual data on self-reported current, former, and never smoking prevalence, data on age at daily tobacco smoking initiation, and time since tobacco smoking cessation. WHS is a nationally representative household survey of adults aged 15 and older, conducted by the Saudi Ministry of Health and based on the World Health Organization’s WHS questionnaires. We used WHS individual data on self-reported current, former, and never tobacco smoking prevalence.

### Tobacco smoking definitions

We briefly describe the tobacco smoking prevalence, age at initiation, and age at cessation definitions used. For more details on the specific survey variables, please see the Supplement (Table S1).

### Prevalence

We categorized individuals in each of the three surveys as never, ever currently, or ever formerly smoking. Never smoking includes those who reported never smoking tobacco products. Ever currently smoking includes those reported ever smoking tobacco products and currently smoking tobacco daily or non-daily (GATS/WHS) or currently smoking tobacco (SHIS). Ever formerly smoking includes those who reported ever smoking tobacco products and who did not currently smoke.

### Age at tobacco smoking initiation

Age at daily smoking initiation was reported by individuals classified as ever currently or formerly smoking in the GATS and SHIS surveys. Age at starting tobacco smoking was reported for the WHS 2019. While the WHS question for age at smoking initiation does not refer to daily smoking, these data were combined with GATS and SHIS to maximize the total sample size.

### Age at tobacco smoking cessation

Time since smoking cessation was reported by individuals who formerly smoked in the GATS survey. Age at tobacco smoking cessation was computed as age at survey minus time since quitting in years. For those reporting quitting less than a year prior to the survey, age at tobacco smoking cessation was set at their current age if quitting less than 6 months prior, and at their current age minus 1 for those quitting more than 6 months prior.

### Model overview

We developed the Kingdom of Saudi Arabia Tobacco Control Policy (TCP) model to simulate the potential impact of a nationwide Tobacco 21 policy in the male population. As smoking rates are extremely low in the female population, preliminary analyses produced unreliable results and precluded model development for the female population.

The model is adapted from the Cancer Intervention and Surveillance Modeling Network (CISNET) TCP model using R version 4.4.1.^13,15^ The model structure is identical to the TCP model but uses different initiation, cessation, and mortality probabilities that are specific to Saudi Arabia. In the model, individuals are born as having never smoked and can transition to current smoking based on annual probabilities of smoking initiation by age (See Supplement, Figure S1). Among those who currently smoke in the model, they can transition to former smoking based on annual probabilities of smoking cessation by age. Regardless of smoking status, individuals can exit the population through annual mortality rates based on age and smoking status. Table 1 summarizes all model inputs and data sources.

**Table 1.**
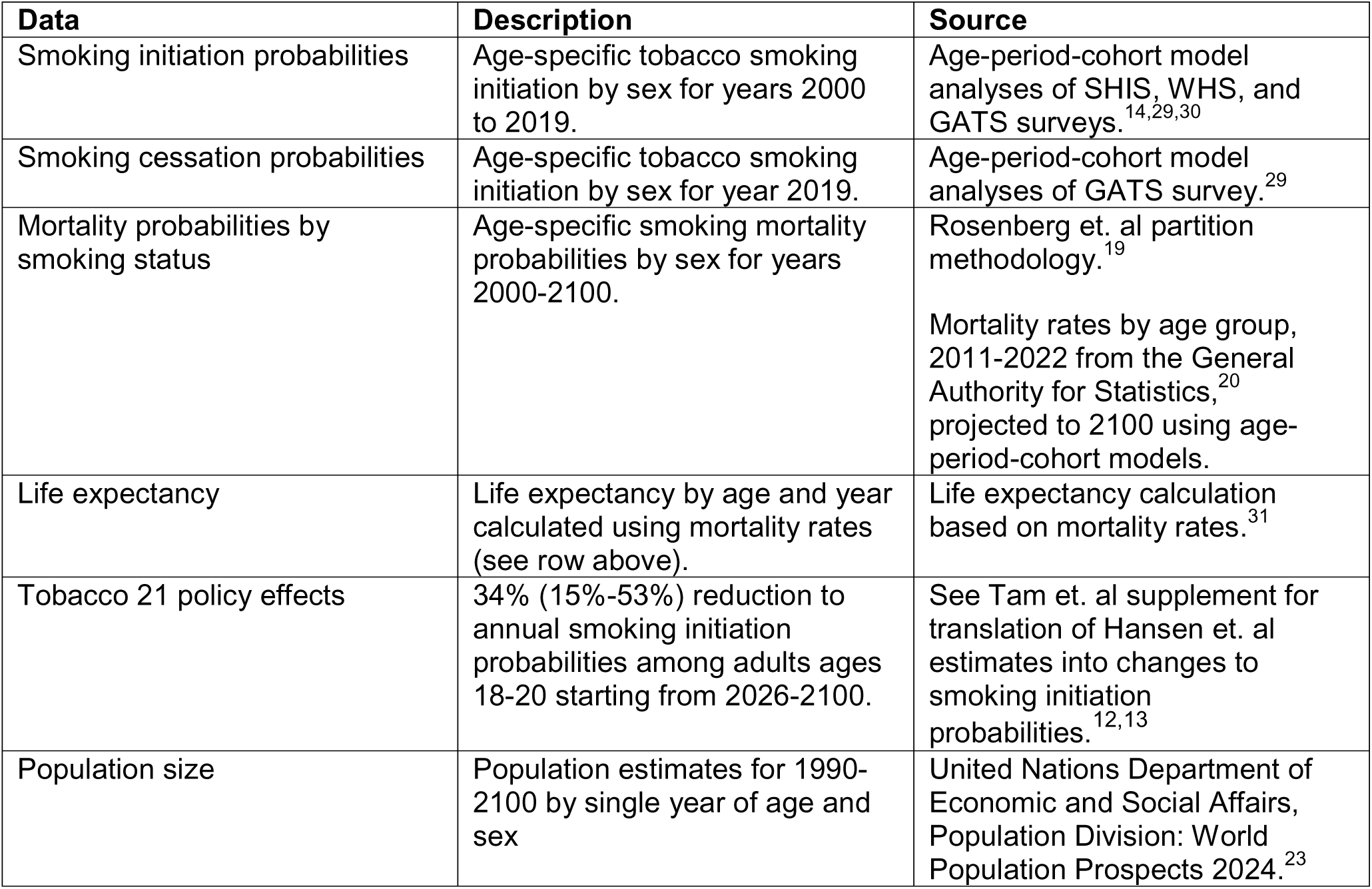
Model inputs and data sources.

### Smoking initiation

We estimated age-specific tobacco smoking initiation probabilities for Saudi males using age-period-cohort models and combined data from the SHIS, WHS, and GATS surveys. If the reported age at initiation was less than 8, it was set at 8, consistent with previous similar analyses by the CISNET consortium for other countries. For each age and birth-cohort or calendar-year represented in the surveys, we then calculated the number of subjects who started to smoke and who had never smoked up to that point. We then fitted age-period-cohort models to the data, similar to previous analyses for smoking initiation for the US and other countries,^16–18^ using the Epi package in R. These analyses provided estimates of smoking initiation probabilities from the year 2000-2019. Initiation is held constant at the 2019 levels for future model projections.

### Smoking cessation

We estimated age-specific tobacco smoking cessation probabilities for Saudi males using age-cohort models and data from the GATS surveys. The analysis was restricted to those classified as ever currently or formerly smoking. If the reported age at smoking cessation was less than 13 years, it was recoded to 13 years, since cessation ages below 13 are considered implausible. For each age and birth-cohort or calendar-year represented in the surveys, we calculated the number of subjects who quit smoking at that age and who had ever smoked to that point. We then fitted age-period-cohort models to the data using the Epi package in R, as above.^16–18^ These provided estimates of smoking cessation probabilities for the year 2019 which are held constant for future model projections.

### Mortality by smoking status

All-cause mortality by age was partitioned by smoking status (never, current, former) using the Rosenberg approach.^19^ In brief, this method combines prevalence estimates for each smoking group from available survey data and relative risks of all-cause mortality associated with current and former smoking for each age group. Mortality rates by age group and sex for the years 2011-2022 from the General Authority for Statistics were used to project annual probabilities death by single year of age through 2100 using age-period-cohort models.^20^ The analysis time horizon extends to 2100 to capture the decades-long time delay between uptake of smoking and subsequent changes in mortality risk. Relative risks of current and former smoking mortality by sex were based on estimates for the US Cancer Prevention Study II.^21^ As in the US TCP model, mortality risk among those who have quit smoking varies by years since quitting; in the first year following cessation, mortality risk begins at the level of someone who currently smokes, and gradually with each year the risk eventually decreases down to the level of someone who has never smoked.^13,15^

### Tobacco 21 policy effects

To apply Tobacco 21 policy effects in the model, we rely on evidence from US quasi-experimental studies that indicate that Tobacco 21 policies reduce smoking initiation for ages 18-20 by 34% (15%-53%).^12,13^ Evidence of similar quality regarding the effects of Tobacco 21 are not available for other settings. Findings were not statistically significant at ages <18, so we assume that the policy has no impact at younger ages. We also assume full implementation and that the policy has no effects on smoking cessation, consistent with the prior modeling analysis.^13,22^ These policy effects are applied as reductions to smoking initiation probabilities for ages 18-20 starting in 2026 continuing through 2100, with the lower and upper bound estimates of 15% and 53% applied as pessimistic and optimistic policy conditions.

### Smoking-attributable mortality

The number of smoking-attributable deaths is calculated for each age according to the following equation using the population size (*P*) for each age (a), the prevalence of current smoking (*prev_cs_*) and former smoking (*prev_fs_*), the age-specific mortality probability for current smoking (*µ_cs_*) or former smoking (*µ_fs_*) individuals:

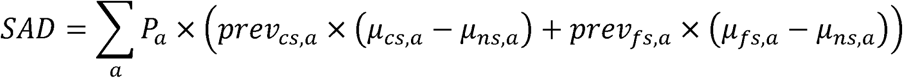

To calculate the number of years of life lost attributed to smoking, each smoking-attributable death is multiplied by the remaining life expectancy of someone who never smoked at that age (*e_ns_*, *a*).

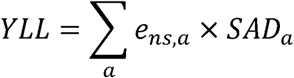

The number of premature deaths averted and life years gained is then determined by taking the difference between the policy and baseline scenarios. Our results do not include any benefits attributed to reduced morbidity or second-hand smoke exposure. Population sizes are based on the United Nations projections (Supplement Figure S2).^23^

### Value of a statistical life cost-benefit analysis

To estimate the monetary value of reducing mortality risk, we multiply the value of a statistical life (VSL) for Saudi Arabia by the number of premature deaths averted. In separate unpublished analyses, the KSA Public Health Authority used a discrete choice experiment to estimate a 2024 VSL range of $1.653 million USD to $5.145 million USD.^24^ Using the fixed exchange rate of 1 USD = 3.75 SAR, this is equivalent to a range of 6.200 to 19.294 million SAR. We use this estimate starting in 2026 and apply an annual discount rate of 3% to projections through 2100.

All model R code, input data, and smoking and mortality estimates will be made available online at https://github.com/mezarafael/KSA_T21 upon publication. To the extent possible, we adhered to the Consolidated Health Economic Evaluation Reporting Standards 2022 (CHEERS 2022) Guidelines.

## RESULTS

Smoking initiation and cessation probabilities by age for males are presented for the year 2026 in Figure 1 and Table S2 (Supplement). As seen in other countries,^25,26^ smoking initiation increases with age, peaks in early adulthood and then decreases. Smoking cessation starts at about 6% in early adolescence but then decreases to ∼2% at age 25. It then increases monotonically with age, a pattern also consistent with those seen in other countries. Figure 1 also shows how the smoking initiation would change under Tobacco 21 policy, assuming the *main* effect estimates. Overall male smoking initiation rates have steadily decreased by calendar year (period) and by birth cohort (See Supplement, Figure S3 and S4).

**Figure 1.**
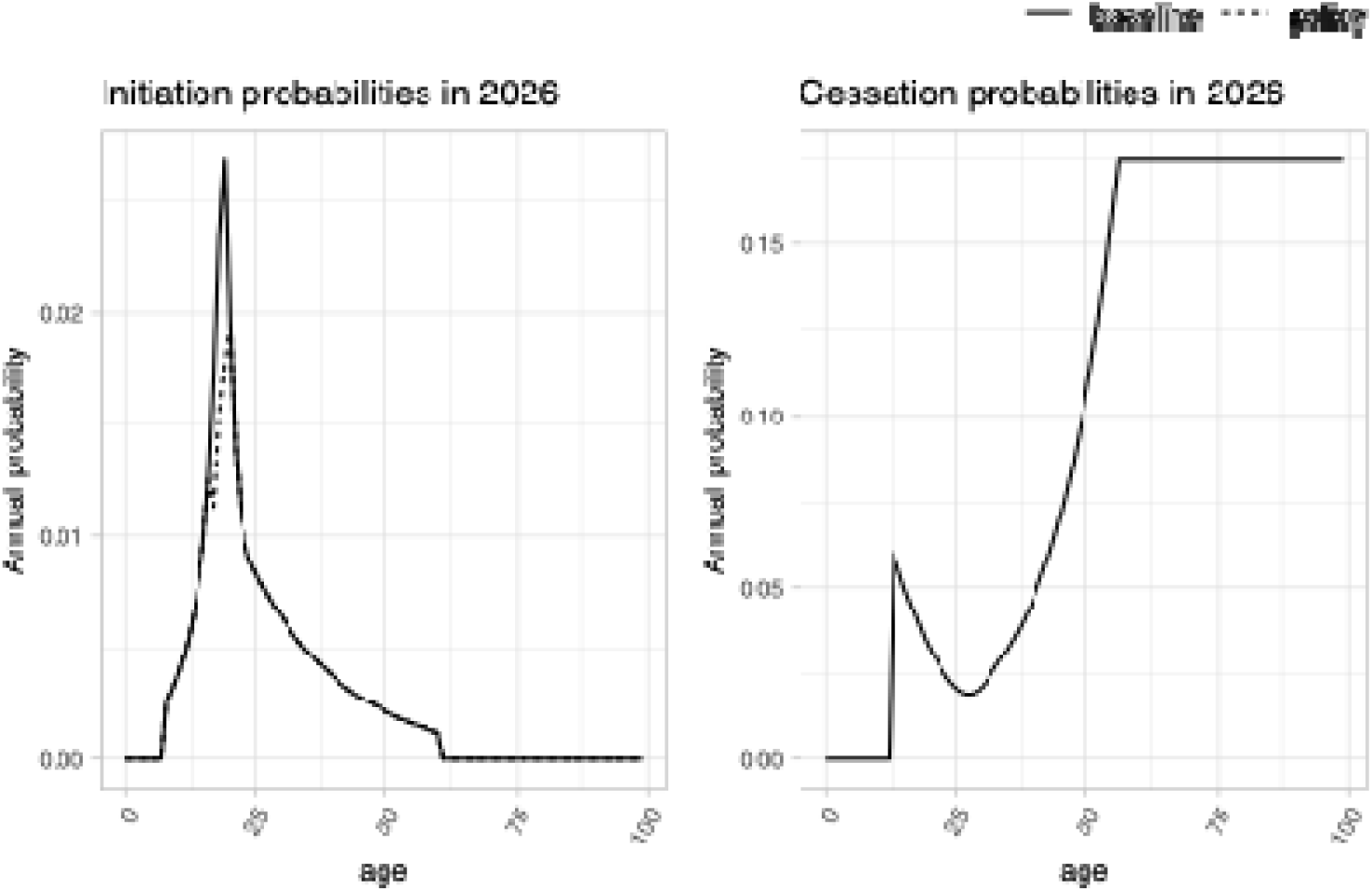
Male smoking initiation and cessation probabilities. Notes: Dashed line represents 34% reduction applied to smoking initiation probabilities for ages 18-20 under the main policy effect scenario. Solid lines represent baseline probabilities.

Age-specific mortality estimates for the year 2026 by smoking status are shown on the actual and log scale in Figure 2. Mortality is high for infants, then decreases to a minimum and then increases in adolescent and youth adulthood years. It then decreases until it settles into a linear increase in the log-scale, a pattern seen in most countries and populations.^27,28^ Mortality for those currently and formerly smoking is higher than that of those who never smoked.

**Figure 2.**
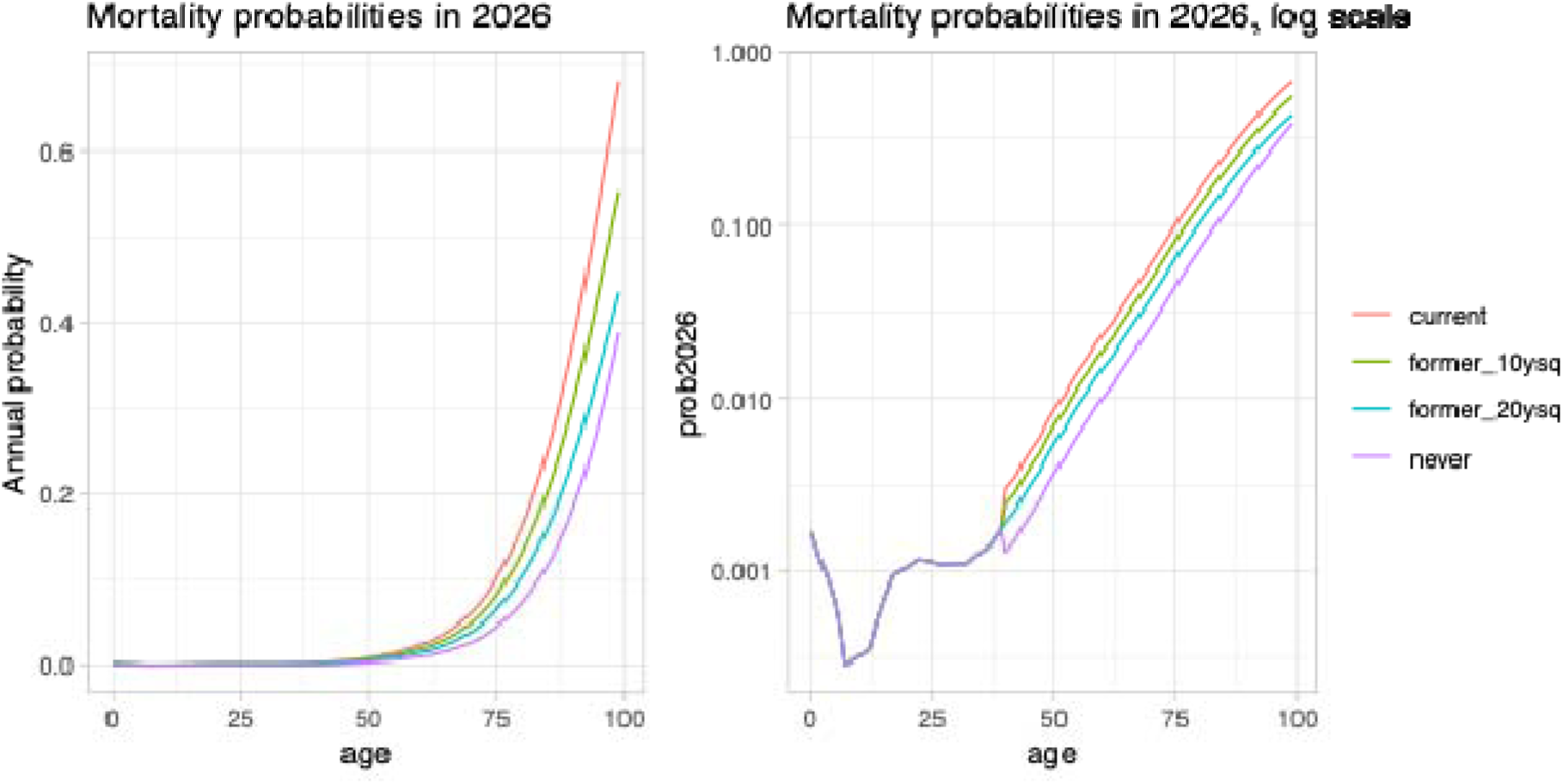
Mortality probabilities by smoking status among males. Notes: Mortality probabilities among those who formerly smoked decrease with time since quitting, starting at the probability of a person who currently smokes and eventually reaching levels of a person who never smoked.. Lines shown for former smoking reflect mortality risk 10 and 20 years since cessation.

Figure 3 shows the model and observed smoking prevalence for males aged 15 or older, with model projections under a status quo scenario. See Supplement Figure S5 for corresponding results for specific age groups. The model estimates align with observed tobacco smoking prevalence in Saudi Arabia from 2013-2019. For 2019, GATS shows a higher smoking prevalence estimate than WHS, with the model being closer to WHS. The model reproduces observed prevalence well for younger ages, but underestimates smoking at older ages. These baseline projections assume that smoking initiation and cessation would remain at the value estimated for 2019. Under these assumptions, the model projects that smoking prevalence would decrease to 10.2% by 2100.

**Figure 3.**
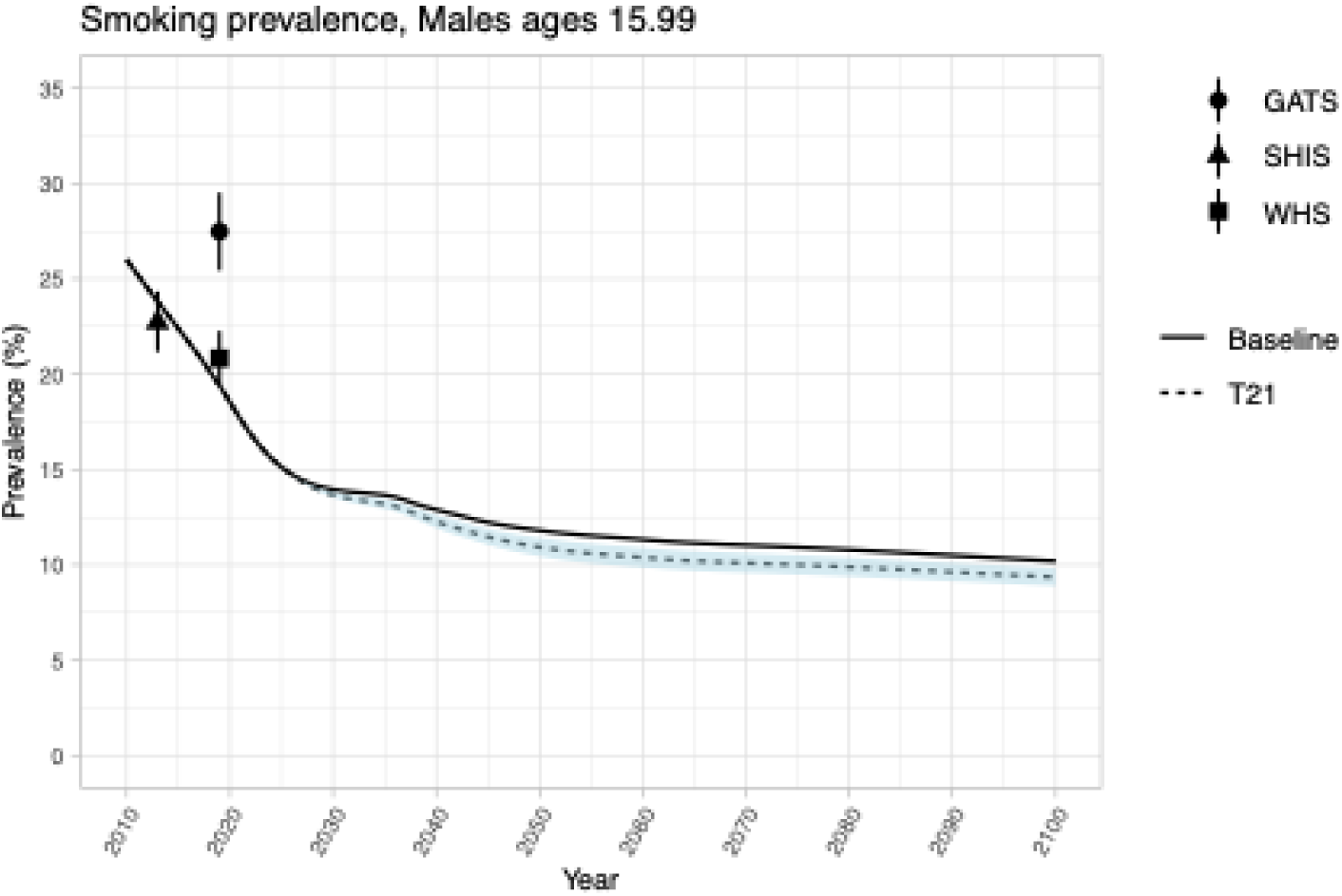
Smoking prevalence 2010-2100, males ages 15+. Notes: Baseline scenario maintains 2019 initiation probabilities going forward. The main Tobacco 21 scenario applies a 34% decrease to initiation probabilities for ages 18-20; the blue ribbon displays 15% to 53% corresponding decreases in initiation, reflecting the range of policy effect estimates.

Implementation of Tobacco 21 would result in slight decreases in tobacco smoking prevalence and corresponding smoking-attributable deaths. By 2100, smoking prevalence would decrease from 10.2% to 9.4% (8.9%, 9.8%) in the main (upper, lower) policy effect scenarios (Figure 3). While modest, these reductions would eventually translate into nearly 5000 (2200, 7800) premature deaths averted with up to 155000 (69000, 241000) life years gained from 2026-2100, respectively (Figure 4).

**Figure 4.**
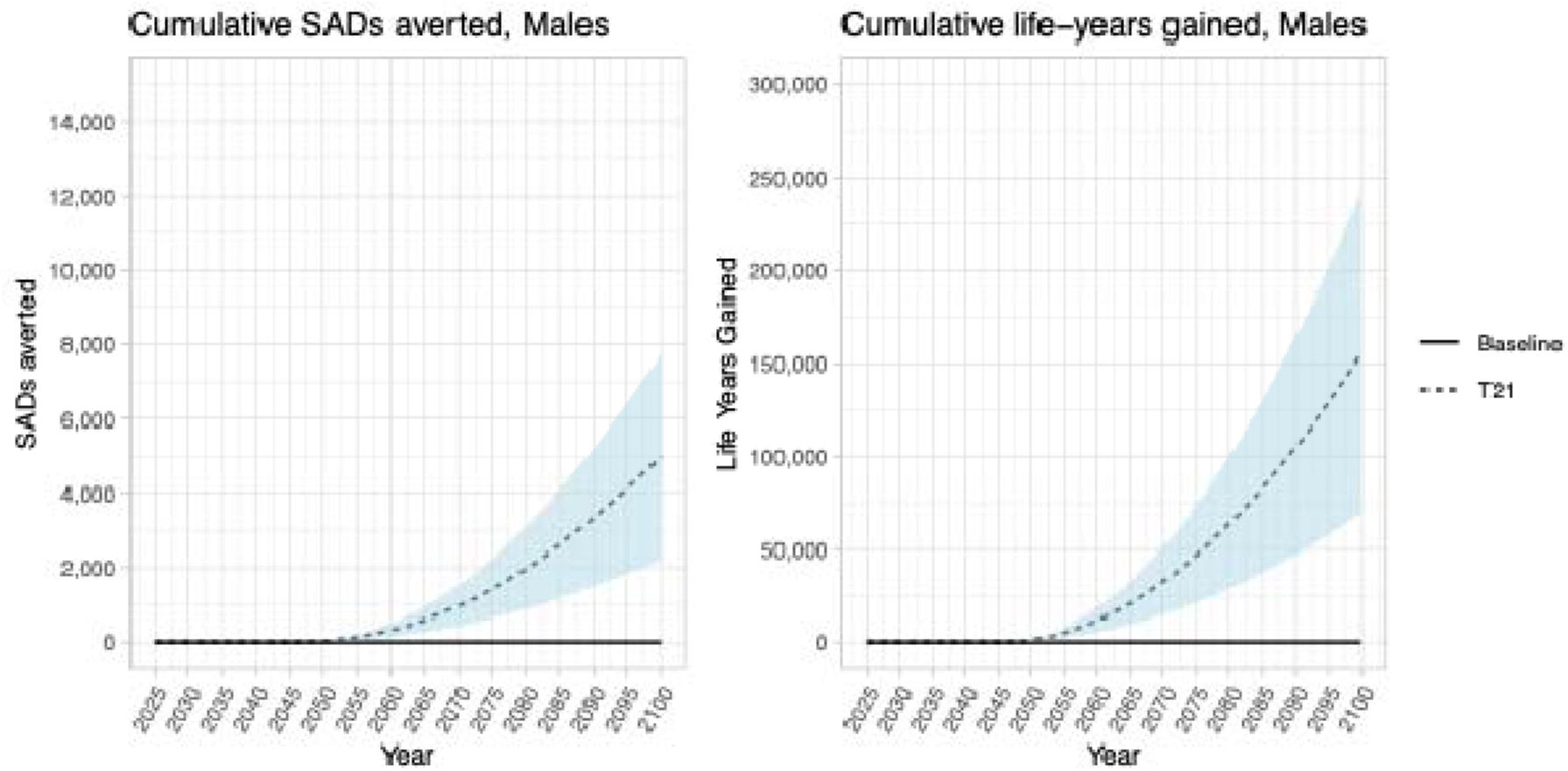
Cumulative premature deaths averted and life-years gained, 2026-2100. Notes: Baseline scenario maintains 2019 initiation probabilities going forward. The main Tobacco 21 scenario applies a 34% decrease to initiation probabilities for ages 18-20; the blue ribbon displays 15% to 53% corresponding decreases in initiation, reflecting the range of policy effect estimates. SADs = smoking-attributable deaths.

Assuming a VSL of $1.653 million USD, the total expected economic benefit is $1.67 ($0.74, $2.60) billion USD, equivalent to 6.25 (2.78, 9.74) billion SAR. Assuming a VSL of $5.145 million USD, this increases the expected benefit to $5.19 ($2.31, $8.09) billion USD, or 19.45 (8.66, 30.32) billion SAR. See Supplement Table S3 for details.

## DISCUSSION

This is the first study to evaluate the potential impact of a Tobacco 21 policy in Saudi Arabia using a simulation model. Through the development of the KSA TCP model, we also present the first estimates of smoking initiation and cessation probabilities in the male Saudi population as well as mortality rates by age, year, and smoking status. We demonstrate that substantial mortality reductions would result from a nationwide law that raises the minimum age of tobacco purchase to 21. Timely implementation would support the KSA in its goals to reduce non-communicable disease and death; however, even under best-case conditions, a Tobacco 21 policy alone will not bring smoking prevalence down to the KSA vision of 9% by 2030; efforts to prevent smoking among young people would need to be combined with interventions that considerably increase smoking cessation among older adults.

There are several important limitations to this study. Regrettably, real-world data on the effects of Tobacco 21 are not available for the KSA or in similarly situated countries. The policy effects evidence used in the model represents the best available data, obtained through quasi-experimental methods, but may not generalize to the Middle East or to other countries with high levels of non-cigarette tobacco smoking. Although other countries have implemented Tobacco 21 policies, they do not have quasi-experimental estimates of policy impacts that can be used in our model. Because the data sources used in the model reflect any smoked tobacco, not just cigarettes, we are unable to disaggregate outcomes by type of tobacco smoking (cigarettes vs. shisha). The smoking initiation and cessation parameters used in the model are based on three data sources with only two calendar years to draw from: 2013 and 2019. The limited historical information makes it difficult to evaluate trends over time and how they may change going into the future. However, even if our model underpredicts or overpredicts smoking prevalence under a baseline scenario, the overall qualitative findings and relative differences under our policy scenarios would still be quite similar. Future trends in underlying smoking initiation and cessation remain subject to uncertainty. We do not include reductions to smoking-related morbidity in this analysis, so our estimates of net health benefit are likely underestimated. This could be an avenue for future research.

This work also raises questions about the consistency and comparability of tobacco measures in national survey data sources. The three surveys used in this analysis differ in design and sampling methods, but without multiple years of data available from the same survey, they are the best available sources for information on adult tobacco smoking in the country. The GATS 2019 data produce higher smoking prevalence estimates compared to the WHS 2019 across all age groups. If evaluating only the SHIS and GATS data, smoking prevalence appears to have increased from 2013 to 2019. However, when evaluating the SHIS and WHS data, their estimates suggest that smoking prevalence has decreased over this period. Based on the estimated parameters and our assumption that initiation and cessation probabilities remain at their 2019 levels going forward, our model projects a slight decrease in prevalence over time. Additional waves of data, using a consistent methodology and sampling, are needed to determine whether smoking rates will continue to decline, plateau, or worsen.

Finally, more data is needed to understand tobacco smoking among female Saudis. Our method of estimating smoking cessation relies on the availability of sufficient numbers of people who have quit smoking in the surveys. In preliminary analyses, we found cessation patterns by age to be implausible due to the small sample size. As a result, we were unable to include females in the model. Although tobacco smoking rates among females are much lower than for males, improved monitoring and surveillance would provide further confidence that female smoking rates remain low in the future.

## CONCLUSION

This study quantifies the potential long-term benefits of a Tobacco 21 policy in Saudi Arabia for the male population. We show that a nationwide Tobacco 21 policy would reduce smoking initiation, smoking prevalence, and smoking-related mortality. Overall smoking prevalence among males ages 15+ would decline, and nearly 5000 premature deaths would be averted with up to 155,000 life years gained from 2026-2100. A Tobacco 21 policy would benefit future generations of young people in the KSA by reducing their risk for heart disease, stroke, and cancer, currently the leading causes of death in the nation.

## Data Availability

Data produced in the present study are available upon reasonable request to the authors. All model R code, input data, and smoking and mortality estimates will be made publicly available online upon publication in a peer-reviewed journal.

## CREDIT AUTHORSHIP STATEMENT

**Conceptualization**: Jamie Tam, Rafael Meza, Taghreed Mohammed Alghaith, Mohammed Adeeb Shahin, Mariam M. Hamza, Reem Alsukait

**Methodology**: Jamie Tam, Rafael Meza, Abdulmohsen H. Al-Zalabani, Sarah Monshi

**Software**: Jamie Tam, Rafael Meza

**Validation**: Jamie Tam, Rafael Meza

**Formal analysis**: Jamie Tam, Rafael Meza

**Investigation**: Jamie Tam, Rafael Meza

**Resources**: Ahmed Ali Yaacoub, Fatmah Mohammed Aldhaher, Abdulmohsen H. Al-Zalabani, Sarah Monshi, Mohammed Abdullah Aljabri

**Data Curation**: Jamie Tam, Rafael Meza

**Writing – Original Draft**: Jamie Tam, Rafael Meza

**Writing – Review & Editing**: Jamie Tam, Rafael Meza, Mohammed Abdullah Aljabri, Abdulmohsen Hamdan Awwad, Sarah Monshi, Mariam M. Hamza Reem Alsukait, Volkan Cetinkaya

**Visualization**: Jamie Tam, Rafael Meza

**Supervision**: Taghreed Mohammed Alghaith, Volkan Cetinkaya

**Project Administration**: Mariam M. Hamza, Reem Alsukait, Fatmah Mohammed Aldhaher

**Funding acquisition**: Mariam M. Hamza, Reem Alsukait, Volkan Cetinkaya

