## Supplement for "Projected impact of a national Tobacco 21 policy in the Kingdom of Saudi Arabia"

**Table S1. Tobacco smoking variable definitions across surveys**

| | GATS 2019 | SHIS 2013 | WHS 2019 |
|---|---|---|---|
| Ever smoking | Currently smoke tobacco on a daily or less than daily basis (*B01 = 1 or 2*)<br>Or<br>Have smoked tobacco in the past daily or less than daily (*B03 = 1 or 2*) | Ever smoked any tobacco products such as cigarettes, cigars, pipes, or Shisha<br>*Tobacco_Smoker_Ever=1* | Ever smoked any tobacco products (cigarettes, waterpipes or cigars) even one time<br><br>*A3001 = 1* |
| Currently smoking | Currently smoke tobacco on a daily or less than daily basis (*B01 = 1 or 2*) | Currently smoke any tobacco products such as cigarettes, cigars, pipes, or Shisha<br>*Tobacco_Smoker_Ever=1*<br>*& Tobacco_Smoker_Current=1* | Currently smoke any tobacco products on a daily basis or less than daily<br><br>*A3003 = 1 or 2* |
| Formerly smoking | Have smoked tobacco in the past daily or less than daily (*B03 = 1 or 2*) | Formerly smoke any tobacco products such as cigarettes, cigars, pipes, or Shisha<br>*Tobacco_Smoker_Ever=1*<br>*& Tobacco_Smoker_Current=0* | Formerly smoked any tobacco products (cigarettes, waterpipes or cigars) even one time<br><br>*A3001 = 1 &*<br>*A3003 = 3* |
| Tobacco smoking initiation | Age at starting smoking tobacco daily<br>*If B01=1, B04*<br>*If B01=2 & B02=1, B08*<br>*If B01=3 & B03=1, B11* | Age at starting smoking daily<br><br>*Tobaco_Age_Startage* | Age at starting smoking tobacco products<br><br>*A3002* |
| Tobacco smoking cessation | Age at stopping smoking<br>Years since stopping: *Age at survey – B13years*<br>Months since stopping>6: *Age at survey minus 1*<br>Months since stopping<6=: *Age at survey* | N/A | N/A |

### Figure S1. Model diagram

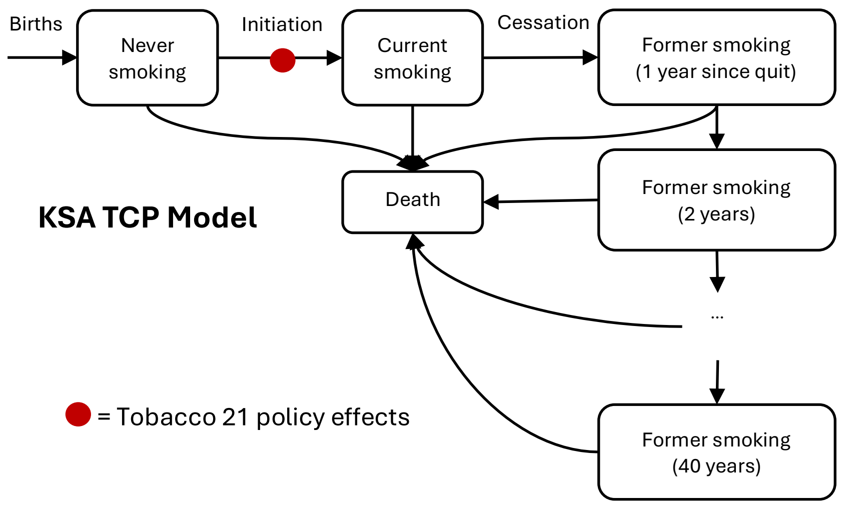


Notes: Adapted from the US Tobacco Control Policy (TCP) Model.[1,2]

### Figure S2. Annual male population in Saudi Arabia, United Nations projections

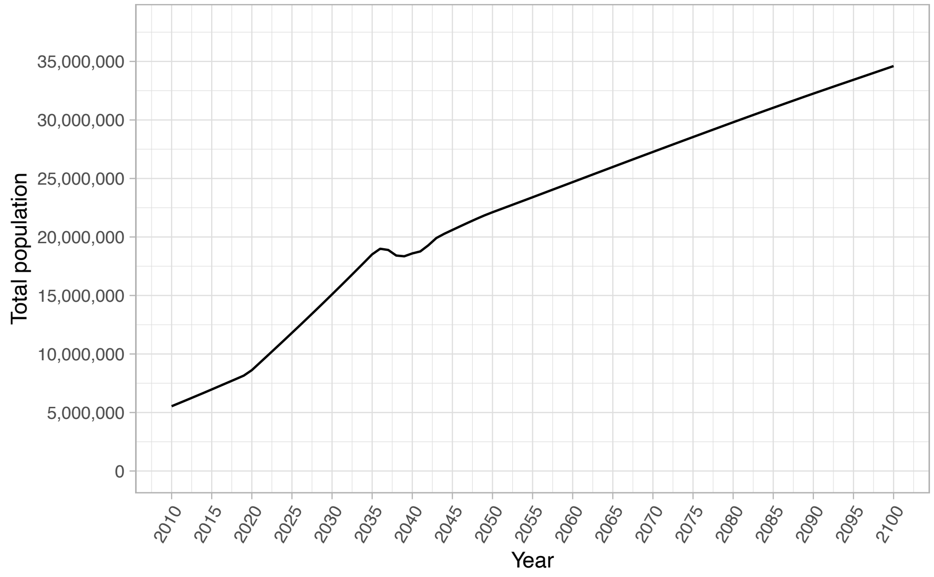


Notes: Population projections from the United Nations Department of Economic and Social Affairs, Population Division: World Population Prospects 2024.[3]

**Table S2. Male smoking initiation and cessation probabilities under baseline scenario, 2019-2100**

| Age | Initiation | Cessation | Age | Initiation | Cessation | Age | Initiation | Cessation |
|---|---|---|---|---|---|---|---|---|
| 0 | 0.0000 | 0.0000 | 35 | 0.0048 | 0.0325 | 70 | 0.0000 | 0.1746 |
| 1 | 0.0000 | 0.0000 | 36 | 0.0046 | 0.0351 | 71 | 0.0000 | 0.1746 |
| 2 | 0.0000 | 0.0000 | 37 | 0.0043 | 0.0378 | 72 | 0.0000 | 0.1746 |
| 3 | 0.0000 | 0.0000 | 38 | 0.0041 | 0.0409 | 73 | 0.0000 | 0.1746 |
| 4 | 0.0000 | 0.0000 | 39 | 0.0039 | 0.0442 | 74 | 0.0000 | 0.1746 |
| 5 | 0.0000 | 0.0000 | 40 | 0.0037 | 0.0478 | 75 | 0.0000 | 0.1746 |
| 6 | 0.0000 | 0.0000 | 41 | 0.0035 | 0.0517 | 76 | 0.0000 | 0.1746 |
| 7 | 0.0000 | 0.0000 | 42 | 0.0033 | 0.0560 | 77 | 0.0000 | 0.1746 |
| 8 | 0.0026 | 0.0000 | 43 | 0.0031 | 0.0607 | 78 | 0.0000 | 0.1746 |
| 9 | 0.0032 | 0.0000 | 44 | 0.0029 | 0.0659 | 79 | 0.0000 | 0.1746 |
| 10 | 0.0038 | 0.0000 | 45 | 0.0028 | 0.0714 | 80 | 0.0000 | 0.1746 |
| 11 | 0.0045 | 0.0000 | 46 | 0.0026 | 0.0775 | 81 | 0.0000 | 0.1746 |
| 12 | 0.0054 | 0.0000 | 47 | 0.0025 | 0.0840 | 82 | 0.0000 | 0.1746 |
| 13 | 0.0065 | 0.0603 | 48 | 0.0024 | 0.0918 | 83 | 0.0000 | 0.1746 |
| 14 | 0.0078 | 0.0551 | 49 | 0.0023 | 0.0995 | 84 | 0.0000 | 0.1746 |
| 15 | 0.0096 | 0.0503 | 50 | 0.0021 | 0.1072 | 85 | 0.0000 | 0.1746 |
| 16 | 0.0125 | 0.0465 | 51 | 0.0020 | 0.1163 | 86 | 0.0000 | 0.1746 |
| 17 | 0.0171 | 0.0427 | 52 | 0.0019 | 0.1262 | 87 | 0.0000 | 0.1746 |
| 18 | 0.0234 | 0.0389 | 53 | 0.0018 | 0.1369 | 88 | 0.0000 | 0.1746 |
| 19 | 0.0268 | 0.0351 | 54 | 0.0017 | 0.1495 | 89 | 0.0000 | 0.1746 |
| 20 | 0.0194 | 0.0321 | 55 | 0.0016 | 0.1620 | 90 | 0.0000 | 0.1746 |
| 21 | 0.0139 | 0.0293 | 56 | 0.0015 | 0.1746 | 91 | 0.0000 | 0.1746 |
| 22 | 0.0111 | 0.0268 | 57 | 0.0014 | 0.1746 | 92 | 0.0000 | 0.1746 |
| 23 | 0.0096 | 0.0245 | 58 | 0.0014 | 0.1746 | 93 | 0.0000 | 0.1746 |
| 24 | 0.0088 | 0.0224 | 59 | 0.0013 | 0.1746 | 94 | 0.0000 | 0.1746 |
| 25 | 0.0083 | 0.0208 | 60 | 0.0012 | 0.1746 | 95 | 0.0000 | 0.1746 |
| 26 | 0.0079 | 0.0197 | 61 | 0.0000 | 0.1746 | 96 | 0.0000 | 0.1746 |
| 27 | 0.0075 | 0.0191 | 62 | 0.0000 | 0.1746 | 97 | 0.0000 | 0.1746 |
| 28 | 0.0071 | 0.0192 | 63 | 0.0000 | 0.1746 | 98 | 0.0000 | 0.1746 |
| 29 | 0.0067 | 0.0200 | 64 | 0.0000 | 0.1746 | 99 | 0.0000 | 0.1746 |
| 30 | 0.0063 | 0.0214 | 65 | 0.0000 | 0.1746 | | | |
| 31 | 0.0060 | 0.0234 | 66 | 0.0000 | 0.1746 | | | |
| 32 | 0.0057 | 0.0256 | 67 | 0.0000 | 0.1746 | | | |
| 33 | 0.0054 | 0.0279 | 68 | 0.0000 | 0.1746 | | | |
| 34 | 0.0051 | 0.0302 | 69 | 0.0000 | 0.1746 | | | |

### Figure S3. Male smoking initiation rates by calendar year

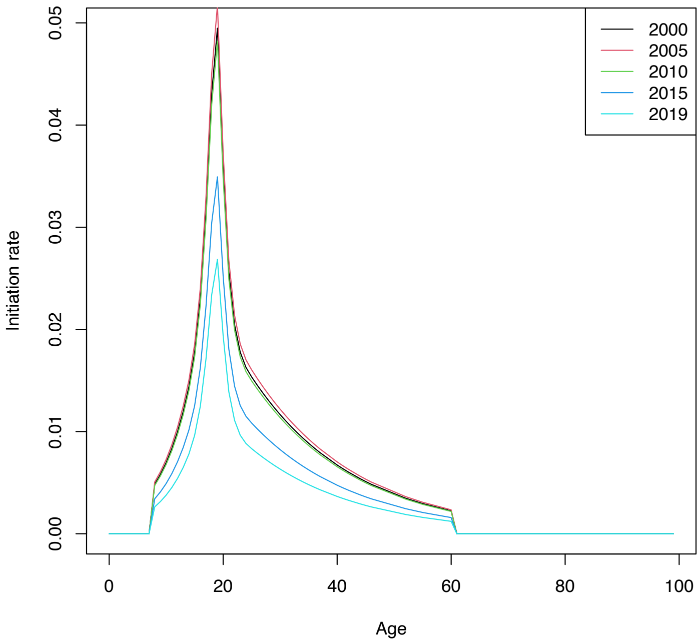


### Figure S4. Male smoking initiation rates by birth cohort

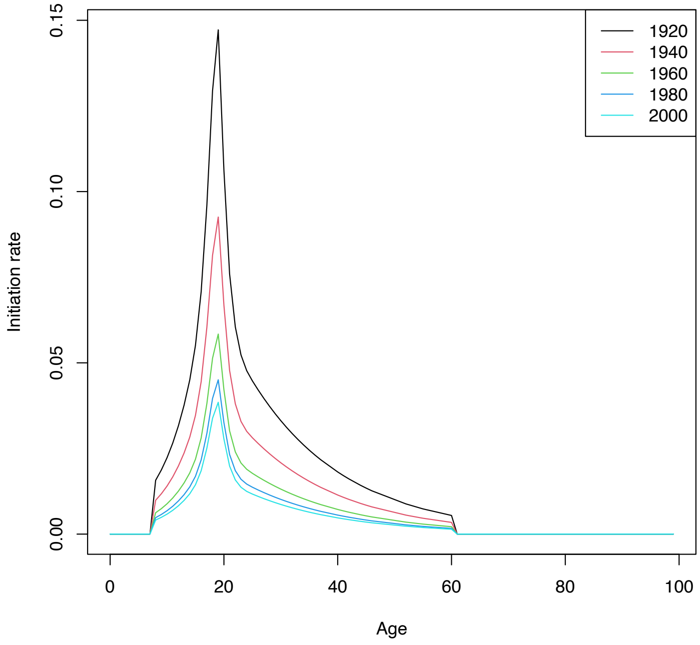

**Figure S5. Smoking prevalence by age group, males ages 15+, 2010-2100**

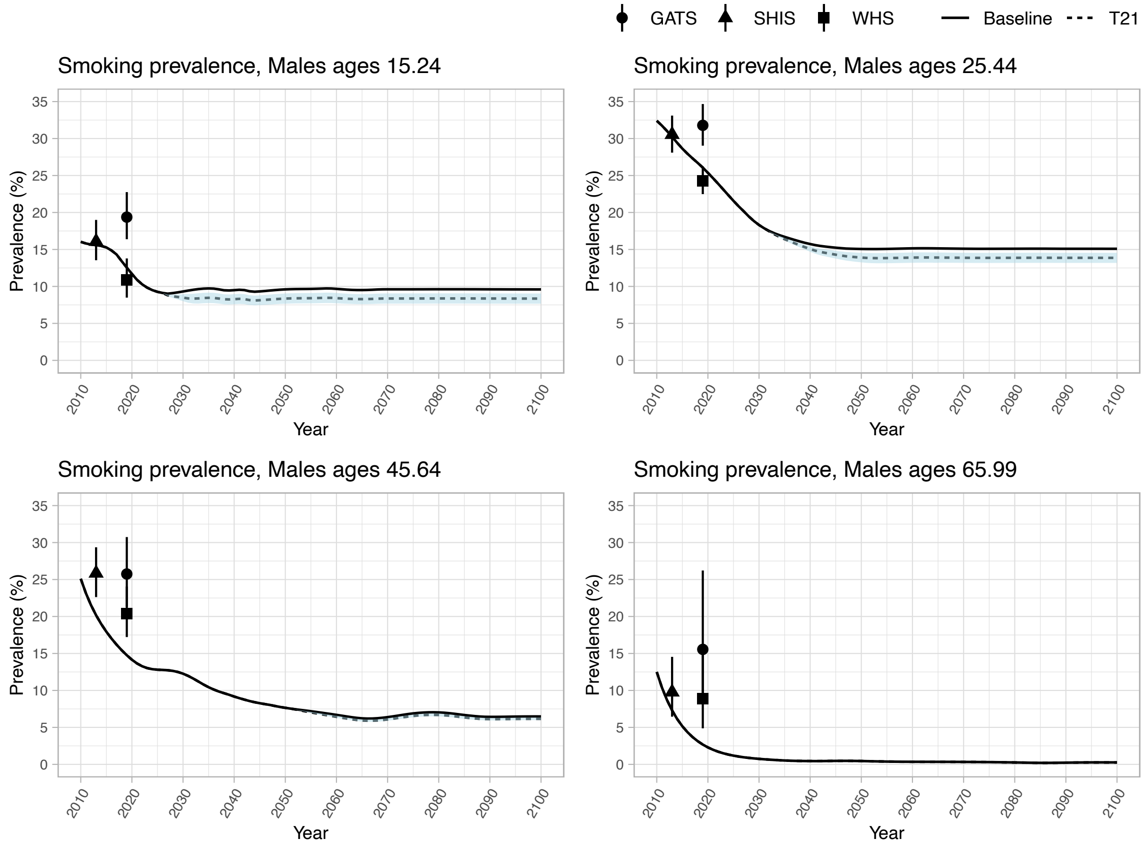


Notes: Baseline scenario maintains 2019 initiation probabilities going forward. The main Tobacco 21 scenario applies a 34% decrease to initiation probabilities for ages 18-20; the blue ribbon displays 15% to 53% corresponding decreases in initiation, reflecting the range of policy effect estimates.

**Table S3. Value of statistical life cost benefit estimation**

| Year | Premature deaths averted | Benefit (millions USD) | Discount_weights | Discounted benefit (millions USD) | Discounted benefit (millions SAR) |
|---|---|---|---|---|---|
| 2026 | 0 | 0 to 0 | 1.00 | 0 to 0 | 0 to 0 |
| 2027 | 0 | 0 to 0 | 0.97 | 0 to 0 | 0 to 0 |
| 2028 | 0 | 0 to 0 | 0.94 | 0 to 0 | 0 to 0 |
| 2029 | 0 | 0 to 0 | 0.92 | 0 to 0 | 0 to 0 |
| 2030 | 0 | 0 to 0 | 0.89 | 0 to 0 | 0 to 0 |
| 2031 | 0 | 0 to 0 | 0.86 | 0 to 0 | 0 to 0 |
| 2032 | 0 | 0 to 0 | 0.84 | 0 to 0 | 0 to 0 |
| 2033 | 0 | 0 to 0 | 0.81 | 0 to 0 | 0 to 0 |
| 2034 | 0 | 0 to 0 | 0.79 | 0 to 0 | 0 to 0 |
| 2035 | 0 | 0 to 0 | 0.77 | 0 to 0 | 0 to 0 |
| 2036 | 0 | 0 to 0 | 0.74 | 0 to 0 | 0 to 0 |
| 2037 | 0 | 0 to 0 | 0.72 | 0 to 0 | 0 to 0 |
| 2038 | 0 | 0 to 0 | 0.70 | 0 to 0 | 0 to 0 |
| 2039 | 0 | 0 to 0 | 0.68 | 0 to 0 | 0 to 0 |
| 2040 | 0 | 0 to 0 | 0.66 | 0 to 0 | 0 to 0 |
| 2041 | 0 | 0 to 0 | 0.64 | 0 to 0 | 0 to 0 |
| 2042 | 0 | 0 to 0 | 0.62 | 0 to 0 | 0 to 0 |
| 2043 | 0 | 0 to 0 | 0.61 | 0 to 0 | 0 to 0 |
| 2044 | 0 | 0 to 0 | 0.59 | 0 to 0 | 0 to 0 |
| 2045 | 0 | 0 to 0 | 0.57 | 0 to 0 | 0 to 0 |
| 2046 | 0 | 0 to 0 | 0.55 | 0 to 0 | 0 to 0 |
| 2047 | 2 | 3 to 8 | 0.54 | 1 to 5 | 6 to 17 |
| 2048 | 5 | 8 to 24 | 0.52 | 4 to 12 | 15 to 46 |
| 2049 | 8 | 14 to 43 | 0.51 | 7 to 22 | 26 to 81 |
| 2050 | 12 | 20 to 61 | 0.49 | 10 to 30 | 36 to 112 |
| 2051 | 15 | 25 to 78 | 0.48 | 12 to 37 | 45 to 140 |
| 2052 | 19 | 31 to 95 | 0.46 | 14 to 44 | 53 to 166 |
| 2053 | 22 | 36 to 112 | 0.45 | 16 to 50 | 61 to 189 |
| 2054 | 25 | 41 to 128 | 0.44 | 18 to 56 | 67 to 210 |
| 2055 | 28 | 46 to 144 | 0.42 | 20 to 61 | 73 to 229 |
| 2056 | 31 | 51 to 159 | 0.41 | 21 to 66 | 79 to 246 |
| 2057 | 34 | 56 to 176 | 0.40 | 23 to 70 | 85 to 263 |
| 2058 | 37 | 62 to 192 | 0.39 | 24 to 75 | 90 to 280 |
| 2059 | 41 | 67 to 208 | 0.38 | 25 to 79 | 95 to 295 |
| 2060 | 44 | 72 to 225 | 0.37 | 27 to 83 | 99 to 310 |
| 2061 | 47 | 78 to 244 | 0.36 | 28 to 87 | 104 to 325 |
| 2062 | 51 | 84 to 262 | 0.35 | 29 to 90 | 109 to 339 |
| 2063 | 55 | 90 to 281 | 0.33 | 30 to 94 | 113 to 353 |
| 2064 | 58 | 96 to 300 | 0.33 | 31 to 98 | 117 to 366 |

| | | | | | |
|---|---|---|---|---|---|
| 2065 | 62 | 102 to 319 | 0.32 | 32 to 101 | 121 to 377 |
| 2066 | 66 | 109 to 338 | 0.31 | 33 to 104 | 125 to 389 |
| 2067 | 69 | 115 to 357 | 0.30 | 34 to 106 | 128 to 399 |
| 2068 | 73 | 121 to 377 | 0.29 | 35 to 109 | 131 to 408 |
| 2069 | 77 | 127 to 396 | 0.28 | 36 to 111 | 134 to 416 |
| 2070 | 81 | 133 to 415 | 0.27 | 36 to 113 | 136 to 424 |
| 2071 | 84 | 139 to 434 | 0.26 | 37 to 115 | 138 to 430 |
| 2072 | 88 | 145 to 453 | 0.26 | 37 to 116 | 140 to 436 |
| 2073 | 92 | 152 to 472 | 0.25 | 38 to 118 | 142 to 441 |
| 2074 | 95 | 158 to 491 | 0.24 | 38 to 119 | 143 to 445 |
| 2075 | 99 | 164 to 510 | 0.23 | 38 to 120 | 144 to 449 |
| 2076 | 103 | 170 to 529 | 0.23 | 39 to 121 | 145 to 453 |
| 2077 | 107 | 176 to 549 | 0.22 | 39 to 121 | 146 to 456 |
| 2078 | 110 | 182 to 568 | 0.22 | 39 to 122 | 147 to 458 |
| 2079 | 114 | 189 to 587 | 0.21 | 39 to 123 | 148 to 460 |
| 2080 | 118 | 195 to 606 | 0.20 | 39 to 123 | 148 to 461 |
| 2081 | 121 | 201 to 625 | 0.20 | 39 to 123 | 148 to 461 |
| 2082 | 125 | 206 to 643 | 0.19 | 39 to 123 | 148 to 460 |
| 2083 | 128 | 212 to 660 | 0.19 | 39 to 122 | 148 to 459 |
| 2084 | 132 | 218 to 678 | 0.18 | 39 to 122 | 147 to 458 |
| 2085 | 135 | 223 to 695 | 0.17 | 39 to 122 | 146 to 456 |
| 2086 | 138 | 229 to 712 | 0.17 | 39 to 121 | 146 to 453 |
| 2087 | 142 | 234 to 728 | 0.16 | 39 to 120 | 145 to 450 |
| 2088 | 145 | 239 to 744 | 0.16 | 38 to 119 | 143 to 446 |
| 2089 | 148 | 244 to 759 | 0.16 | 38 to 118 | 142 to 442 |
| 2090 | 150 | 249 to 774 | 0.15 | 37 to 117 | 141 to 438 |
| 2091 | 153 | 253 to 788 | 0.15 | 37 to 115 | 139 to 433 |
| 2092 | 156 | 257 to 801 | 0.14 | 37 to 114 | 137 to 427 |
| 2093 | 158 | 262 to 814 | 0.14 | 36 to 112 | 135 to 421 |
| 2094 | 161 | 265 to 826 | 0.13 | 36 to 111 | 133 to 415 |
| 2095 | 163 | 269 to 838 | 0.13 | 35 to 109 | 131 to 409 |
| 2096 | 165 | 273 to 850 | 0.13 | 35 to 107 | 129 to 403 |
| 2097 | 168 | 277 to 862 | 0.12 | 34 to 106 | 127 to 396 |
| 2098 | 170 | 281 to 874 | 0.12 | 33 to 104 | 125 to 390 |
| 2099 | 172 | 285 to 886 | 0.12 | 33 to 102 | 123 to 384 |
| 2100 | 174 | 288 to 898 | 0.11 | 32 to 101 | 121 to 378 |
| Total | 4975 | 8223 to 25595 | | 1666 to 5186 | 6248 to 19447 |

Notes: Reflects estimates under the main T21 policy scenario with a 3% discount rate. Estimates are rounded to whole numbers for display purposes. The calculation applies the 2024 KSA value of a statistical life which ranges from 1.653 to 5.145 million USD.
